# Unveiling the Awareness of Private Health Insurance Coverage among Healthcare Professionals in Freetown, Sierra Leone: Insights Extracted from Their Perspectives

**DOI:** 10.64898/2026.06.11.26355471

**Authors:** Sallu Nfagie Kamara, Abraham Isiaka Jimmy, Aiah Lebbie, Okube Tekeste, Lee Presley Gary

**Affiliations:** Department of Pharmacology, College of Medicine and Allied Health Sciences, University of Sierra Leone, Freetown, Siera Leone; Department of Public Health, Faculty of Nursing and Allied Health Sciences, University of Makeni, Makeni, Sierra Leone; Department of Surgery, College of Medicine and Allied Health Sciences,University of Sierra Leone, Freetown, Siera Leone; Office of Faculty Research, University of Makeni, Makeni, Sierra Leone

**Keywords:** Universal Health Coverage, private health insurance, premium payments, insurance coverage, healthcare professionals, Low-and Middle-Income Countries, and Freetown, Sierra Leone

## Abstract

Our study is an assessment of the knowledge, personal coverage, and related determinants of private health insurance as revealed by healthcare professionals in Freetown, the urban capital of Sierra Leone. This study stands as a precursor for Low- and Middle-Income Countries (LMICs), like Sierra Leone, seeking to establish Universal Health Coverage (UHC) to provide healthcare access and coverage through publicly arranged risk pooling, designed to help protect against unmanageable medical costs. In parallel, such countries face significant challenges with achieving sustainable universal coverage due to limited public resources, inefficient allocation systems, uneasy reliance on out-of-pocket payments, and large struggling populations. Our research sheds particular light on how healthcare professionals view their own participation with private healthcare options. A cross-sectional, analytical study was conducted, openly recruiting individuals from various facilities in Freetown. Using the Yamane Formula, a sample size of 109 participants was calculated. STATA 14.0 was used for data analysis. Our findings revealed that 96 (88.9%) participants did not have private health insurance, while 12 (11.1%) did have private coverage. However, 105 (97.2%) reported other modes of health insurance, with only 3 (2.8%) uninsured. Notably, 97.2% expressed willingness to join a private health insurance scheme. Our study found no statistically significant associations between selected indicators (demographic or socioeconomic fac tors) and current insurance coverage among study participants. These results highlight a low prevalence and understanding of private health insurance among healthcare professionals in a representative urban center in Sub-Saharan Africa (SSA), while acknowledging high willingness to enroll. The lack of anysignificant determinants suggests other unexamined factors, such as cost, accessibility, or awareness, capable of influencing the adoption and implementation of a universal health program.

## 1. INTRODUCTION

The current health care financing system in Sierra Leone is unsustainable and poses challenges ranging from increases in out of pocket health (OOP) care expenditures to accessibility problems, particularly in rural areas where living standards are low and health care facilities are scarce.?? Still after more than sixty (60) years of independence, inequalities with access to health care are widely prevalent in Sierra Leone communities. S u c h inequalities in access to health care are related principally to socioeconomic status, geography, and gender, and are compounded by high out-of-pocket expenditures, with more than three-fourths of the increasing financial burden of health care being met by households.?? The rise in health care demand has increased the cost of the health care system to the extent that specialized care is beyond the reach of most individuals. [8]. There is a need to provide a financial shield to health care providers for these reasons. Hence this study seeks to do an assessment among Health Care Professionals (HCP) on private health insurance being one of the obvious healthcare financing mechanism.

### 1.1 Knowledge of Private Health Insurance

Iin a study conducted by Setswe et al. [17], where National Health Insurance (NHI) in South Africa prevails, 49.8% of respondents didn’t know how the NHI functions as a whole. 47.1% of respondents, were aware that all NHI patients will first access primary care (clinic or General Practitioner) level of services. If necessary, the primary healthcare provider would subsequently refer the patient to a hospital or a specialist. This is less than 50.1% who did not know. 48.7% of respondents were unaware that NHI will be expected to work with high level service providers, compared to 46.9% of respondents who were aware that the same standard of care was expected from commercial and public healthcare providers.

### 1.2 Private Health Insurance Coverage

One key tool for improving provider and care level options is private health insurance (PHI). Despite providing coverage for a significant portion of the population, PHI remained??? a key tenet of the healthcare system. Those who elect to be admitted as private patients are covered by private hospital insurance for hospitalization in either private or public hospitals. Many government incentives have also encouraged private health insurance, but policy shifts have favored either private or public financing of the healthcare system [2]. In many low- and middle-income nations, the private sector is the main supplier of primary healthcare for the underprivileged. Both official, or by legally recognized regulatory authorities, and informal, or not recognized by the law, private healthcare providers are possible. For-profit and nonprofit hospitals are included in the category of formal private healthcare providers. For-profit/not-for-profit dichotomy is not so distinct in practice. Informal allopathic ?? providers includes quacks lay health workers, drug sellers and ordinary shop keepers. Public healthcare providers on the other hand, are health facilities built by government and the healthcare workers draw their salaries from government treasuries. This facility type is not intended for profit making [13].

Despite this, studies on disability among senior people have paid much less attention to health insurance coverage than other risk variables [9]WHERE ???. According to Smith and Medalia [18], from their study, the majority of persons (89.6%) had access to health insurance at some time in the year, with private health insurance coverage being more common (66.0%) than government coverage (36.5%). Employer-based insurance, which covered 55.4% of the population, was the most common type of health insurance, followed by Medicaid (19.5%), Medicare (16.0%), direct purchase (14.6%), and military health care (4.5%).

### 1.3 Determinant Associated with Health Insurance Coverage

Health insurance serves as a source of funding for medical costs. While the majority of people WHERE ?? have private health insurance, typically provided by their employers, many more are covered through government-sponsored programs. Some people have no health insurance at all [18]. Changes in the rate of health insurance coverage and the distribution of coverage types over time may be a reflection of societal demographic transitions, economic trends, and policy changes that affect patient access to careThey appear to have a say in decisions regarding health insurance as well. Similar to this, some consumers may not be able to assess whether the advantages of health insurance outweigh its costs due to a lack of information about terms like deductibles or copayments [4]. In middle- and low-income countries, there is a limited but growing corpus of econometric research that has examined household (demographic and economic) and individual (sex, age, income, education) determinants of demand for health insurance. The data indicates that people with greater socioeconomic status are more likely to have health insurance [3].

## 2. METHODOLOGY

### 2.1 Study Design

Our project is a cross-sectional re s earch study and analytic? By design, healthcare workers (professional and technical) in various facilities in Freetown were recruited.?? This study type and design were chosen as the best fit for the focus of the project: to confirm by survey what Healthcare Professionals in Freetown know about the nature and scope of private healthcare insurance. Also, the cross-sectional design is appropriate for studies in which the presence or absence of disease or other health-related variables are determined at a single point in time or time interval which best describes the study at hand.??

### 2.2 Study Location and Population

This study was conducted in various health facilities in Freetown, the Capital of Sierra Leone. It serves as the country’s principal urban, financial, cultural, educational, and political hub. Free town has the largest natural harbor on the African continent, which serves as the economic hub of the city and nation. The population of Freet ownis ??? ? (Reference)

The study population in composed of healthcare workers in selected facilities:(Connaught Teaching Hospital, Pharmacy Board of Sierra Leone, National Medical Supplies Agency, Directorate of Pharmaceutical Service, Malaria Program, and Epidemiology Unit, all in Freetown. in Freetown. The population size of healthcare workers in Freetown is considered to be 487.

### 2.3 Sampling and Sampling Technique

Given the known population size of healthcare workers, the sample size for the study was calculated using the Yamane formula (95% confidence level and 0.09 error margin). The formula for calculating the sample size is given by:

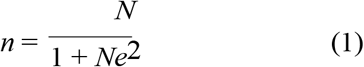

where:

2.3.1 *n*: Sample size

2.3.2*N*: Study population

2.3.3*e*: Margin of error

For a study population *N* = 487 and a margin of error *e* = 0.09, the sample size is calculated as follows:

Therefore, at the confidence level of 95% and the precision rate of 0.09, the sample size for this study is 109. A probability sampling technique was used for this study. Using probability proportion to size sampling, is an approach that is useful when the units are of unequal sizes, and ensures the likelihood of a unit being selected is proportionate to the size of the represented population.

### 2.4 Data Collection

Data for this study was obtained from research questions. For all research questions, quantitative data was extracted, . Respondents were randomly selected using a simple random sampling ?? until the targeted number was reached. Health care workers were given codes based on the proportion determined for the study. The codes were inscribed with the words “yes” or “no” on large cardboard pieces, folded, and put in a bowl. To prevent biases, the cardboards were carefully blended. Randomly selected health care workers were asked to pick up a piece of the cardboard containing the codes one at a time. This went on until the necessary sample size was reached. Those who selected “yes” and provided their full consent were enrolled in the study. On the other side, those who responded “no” were not included in the study. Data coding was performed by designing a questionnaire that assigns a number to each response for data entry. Research assistants were trained how to administer the questionnaire and data collection procedures. Successive study participants were identified by the principal researcher following the inclusion and exclusion criteria. Following the selection of the responders, each study participant was given a personalized explanation of the study in a language they could understand, preferably Krio. To ensure the accuracy and reliability of the questionnaire for data collection, a pre-testing of the tool was done in a facility which was not part of the study. During the pre-testing, it was found that few important questions were not included. Based on that, a review was made to make it more appropriate for data collection.

### 2.5 Data Analysis

The Statistical Package, STATA V.14.0, was used to conduct the statistical analysis. Microsoft Excel was used to enter and clean the data collected for this study. On a 95% confidence level, both descriptive and inferential analysis were performed for each target????. Tables with frequencies, percentages, odds ratios, and coefficients were used to present the results.

## FINDINGS

### Socio-Demographic Characteristics

According to the study, Table 1 shows the socio-demographic characteristics of the respondents, which polled 109 healthcare professionals in Sierra Leone; the majority of them were under 30 years old. The sample is almost evenly divided between Muslims and Christians, with a small male preponderance. Smaller teams are employed by the Pharmacy Board, Malaria Program, Directorate of Pharmaceutical Services, Epidemiology Unit, and National Medical Supplies Agency, but the majority are employed at Connaught Teaching Hospital. The most prevalent occupations are those of pharmacists and nurses, followed by medical doctors, community health officials, and laboratory/pharmacy technicians. Less than half have five to nine years or ten or more years of job experience, while more than half have less than five. A moderate monthly income is earned by the majority, with the greatest group falling into the mid-range income bracket, followed by higher-earning individuals and smaller groups in the lower and upper-middle income range.

**Table 1.** Demographic and Professional Characteristics of Respondents (n=109)

| Variable | Frequency<br>(n=109) | Percentage<br>(%) | Mean | SD |
| --- | --- | --- | --- | --- |
| **Age category of Respondents** |  |  |  |  |
| 30 and Below | 61 | 56.48 | 32.14 | 8.22 |
| 31–35 | 15 | 13.89 |  |  |
| 36–40 | 14 | 12.96 |  |  |
| 41–45 | 10 | 9.26 |  |  |
| Above 45 | 8 | 7.41 |  |  |
| **Gender of Respondent** |  |  |  |  |
| Female | 48 | 44.44 |  |  |
| Male | 60 | 55.56 |  |  |
| **Religion of Respondents** |  |  |  |  |
| Christianity | 51 | 47.22 |  |  |
| Islam | 57 | 52.78 |  |  |
| **Place of work** |  |  |  |  |
| Connaught Teaching Hospital | 64 | 59.26 |  |  |
| Directorate of Pharmaceutical Services | 9 | 8.33 |  |  |
| Epidemiology Unit | 8 | 7.41 |  |  |
| Malaria Program | 10 | 9.26 |  |  |
| National Medical Supplies Agency | 3 | 2.78 |  |  |
| Pharmacy Board of Sierra Leone | 14 | 12.96 |  |  |
| **Professional cadre** |  |  |  |  |
| CHO | 8 | 7.41 |  |  |
| Laboratory/Pharmacy technicians | 25 | 23.15 |  |  |
| Medical Doctor | 7 | 6.48 |  |  |
| Nurse | 31 | 28.70 |  |  |
| Pharmacist | 37 | 34.26 |  |  |
| **Length of years of operation** |  |  |  |  |
| ≤5 years | 56 | 51.85 |  |  |
| 5–9 years | 29 | 26.85 |  |  |
| 10 years and above | 23 | 21.30 |  |  |
| **Monthly Income of Respondents (NLe)** |  |  |  |  |
| 2000 and Below | 19 | 17.59 | 65328.21 | 577673.30 |
| 2001–4000 | 19 | 17.59 |  |  |
| 4001–6000 | 32 | 29.63 |  |  |
| 6001–8000 | 15 | 13.89 |  |  |
| Above 8000 | 23 | 21.30 |  |  |

### Knowledge on Private Health Insurance

Table 2’s findings indicate that while a smaller percentage are unaware of private health insurance, the majority are. Of those who are aware, most believe that private health insurance makes life easier and covers medication cost reimbursements during illness; more than half also acknowledge that it offers free major surgeries, while fewer link it to recompense for unfavourable outcomes. A little more than half of all respondents say they are aware of private health insurance, while the remaining respondents say they a e n?

**Table 2.** Awareness and Perceptions of Private Health Insurance Among Respondents (n=109)

| Variable<br>(n=109) | Frequency | Percentage<br>(%) |
| --- | --- | --- |
| **Know what is a private health insurance** |  |  |
| No | 27 | 25.00 |
| Yes | 81 | 75.00 |
| **Know private health insurance to be as: (Multiple Response)** |  |  |
| Refund of cost of drugs during illness | 68 | 80.95 |
| Insurance makes life easier | 60 | 71.43 |
| Provide free of cost major surgeries | 53 | 58.89 |
| Provide compensation if something bad happens | 31 | 36.90 |
| **Know of any private health insurance** |  |  |
| No | 48 | 44.44 |
| Yes | 60 | 55.56 |

**Figure 1.**
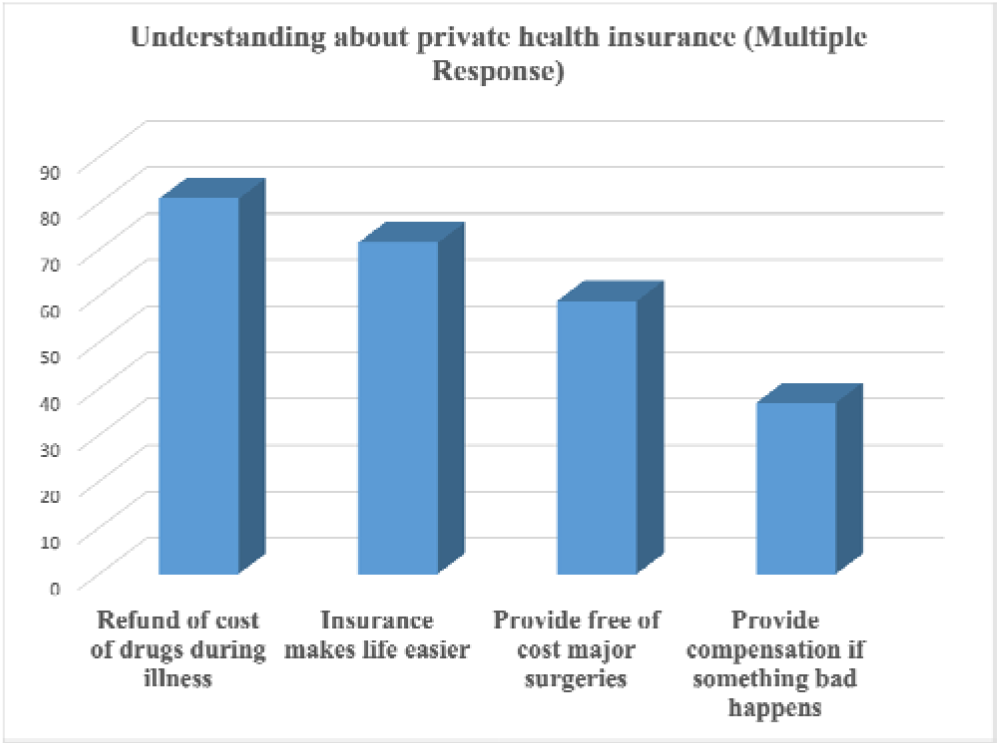
Understanding about private health insurance

### 2.1. Insurance Coverage

According to the study’s findings in Table 3, just a small percentage of respondents had health insurance, while themajority did not. Only a small percentage of respondents reported having insurance at the moment, while the majority are uninsured. Furthermore, while a minority indicate no interest in joining a health insurance program, the clear majority say they would be willing to do so.

**Table 3.**
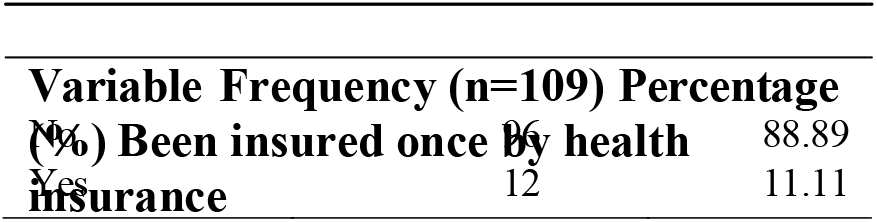
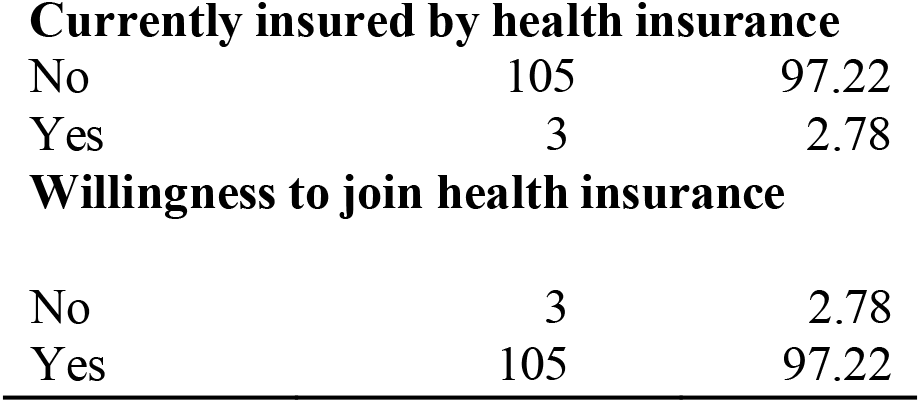
Health Insurance Status and Willingness Among Respondents (n=109)

**Table 4.**
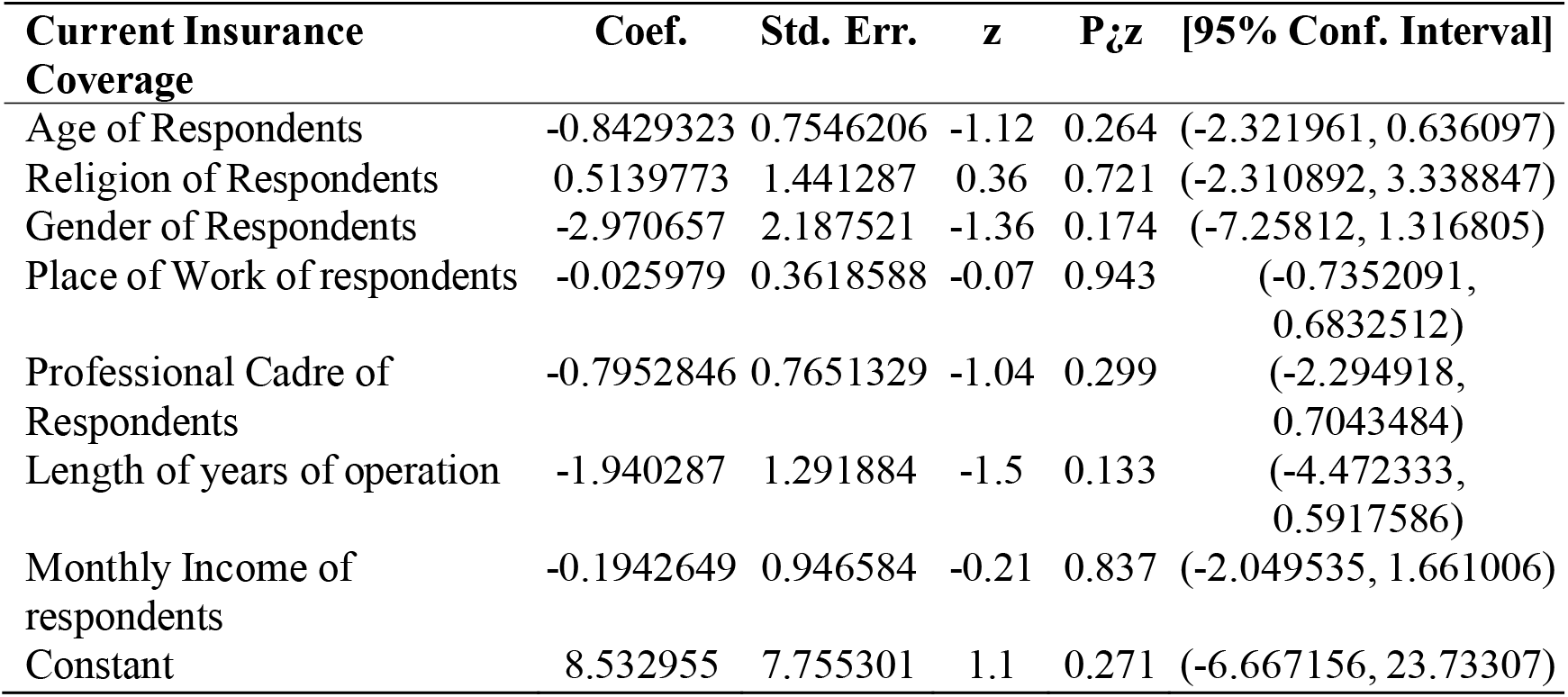
Factors associated with Current Insurance Coverage.

| Current Insurance Coverage | Coef. | Std. Err. | z | P>z | [95% Conf. Interval] |
| --- | --- | --- | --- | --- | --- |
| Age of Respondents | -0.8429323 | 0.7546206 | -1.12 | 0.264 | (-2.321961, 0.636097) |
| Religion of Respondents | 0.5139773 | 1.441287 | 0.36 | 0.721 | (-2.310892, 3.338847) |
| Gender of Respondents | -2.970657 | 2.187521 | -1.36 | 0.174 | (-7.25812, 1.316805) |
| Place of Work of respondents | -0.025979 | 0.3618588 | -0.07 | 0.943 | (-0.7352091, 0.6832512) |
| Professional Cadre of Respondents | -0.7952846 | 0.7651329 | -1.04 | 0.299 | (-2.294918, 0.7043484) |
| Length of years of operation | -1.940287 | 1.291884 | -1.5 | 0.133 | (-4.472333, 0.5917586) |
| Monthly Income of respondents | -0.1942649 | 0.946584 | -0.21 | 0.837 | (-2.049535, 1.661006) |
| Constant | 8.532955 | 7.755301 | 1.1 | 0.271 | (-6.667156, 23.73307) |

**Figure 2.**
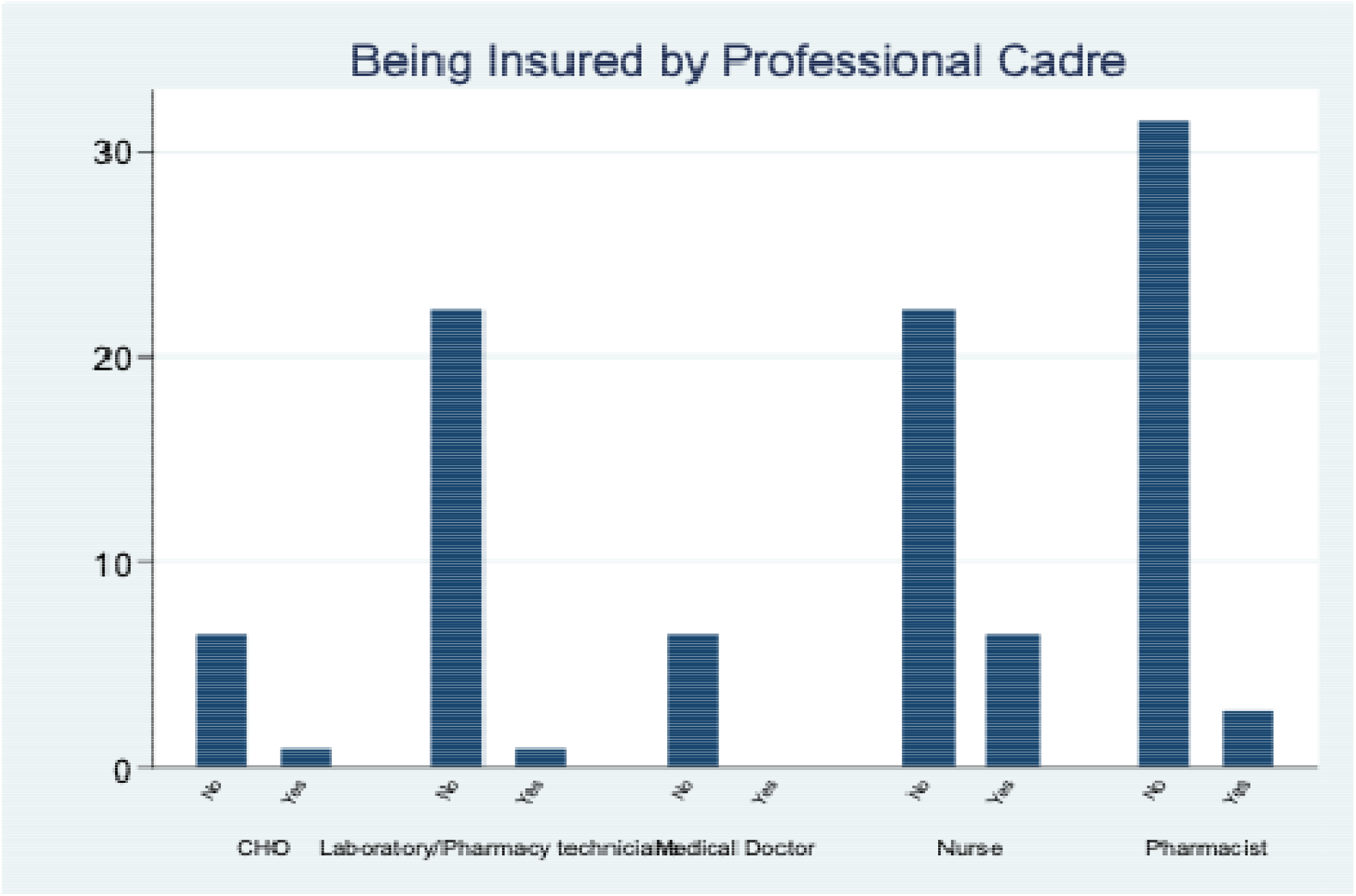
Insurance status by Professional Cadre

Some CHOs are being insured but some are not, majority of the laboratory / pharmacy technicians are not health insured only a little are insured. Almost all medical doctors are not health insured, few of the nurses are insured while majority are not. Lastly, most pharmacist are not health insured but a few are insured.

**Figure 3.**
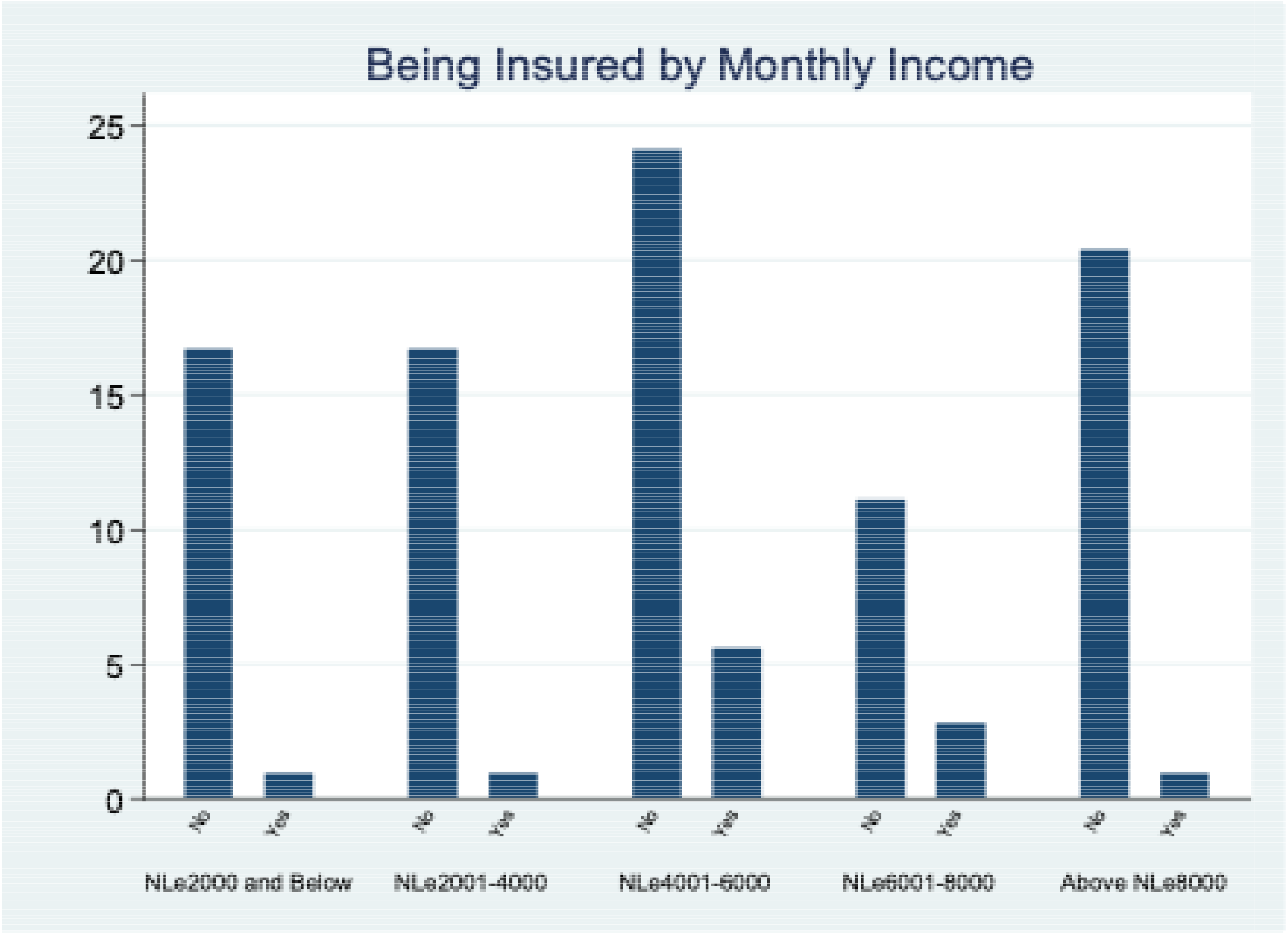
Being Insured by Monthly Income

Most respondent with NLe 2000 and below are not insured only a little are insured, those with NLe 2001-4000 are also not health insured only a few are insured. Participants who receives NLe 4001-6000 are not being insured the same if for respondents that receives NLe 6001-8000 and NLe 8000 above only a few of them said yes they are health insured.

**Figure 4.**
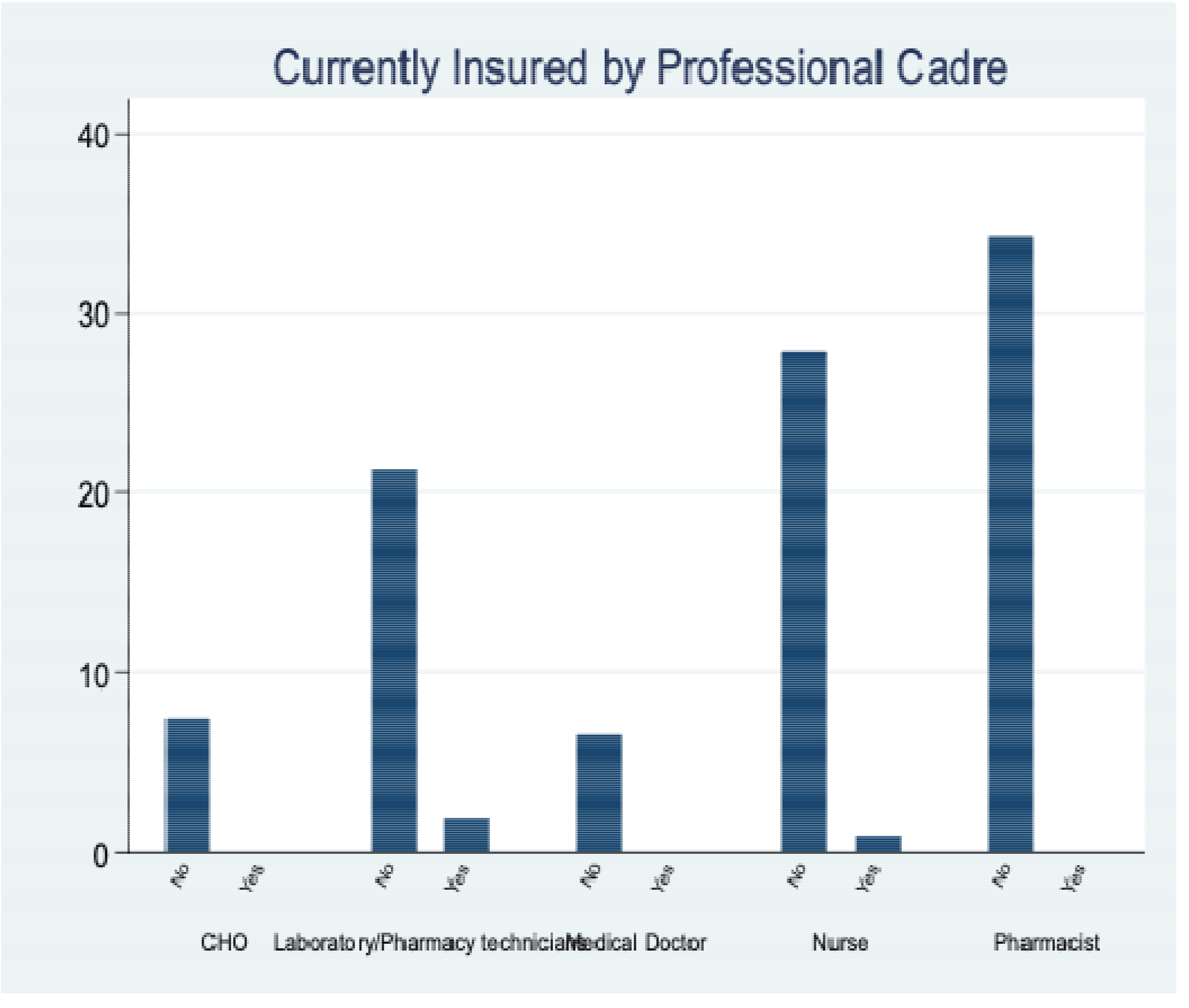
Currently health insured by Professional cadre

Currently, no CHO are health insured, pharmacists are also not currently health insured the same is for medical doctor. In the order caders like Nurces and Laboratories, only few of the are currently insured.

**Figure 5.**
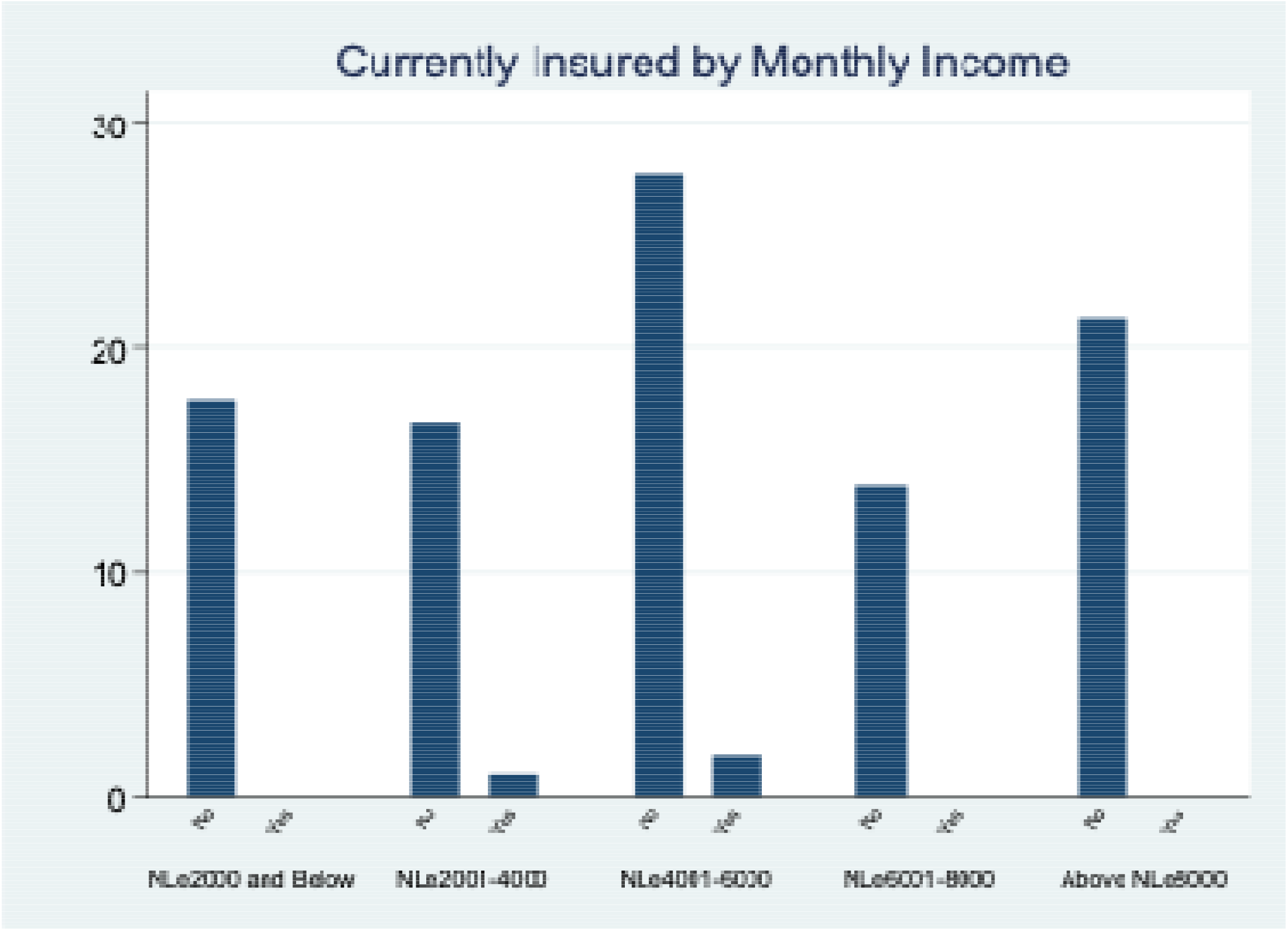
Currently insured by Monthly income

### 2.2. Factors associated with Current Insurance Coverage

From the Regression analysis to identify determinants associated with the current insurance coverage. From the research it shows that none of the variables are not statistically significant.

## 3. DISCUSSION

The first objective of this study was to evaluate the knowledge of healthcare workers regarding private health insurance. Findings indicate that a significant portion of respondents are familiar with the concept??, while a smaller group remains unaware. Among those aware, the majority perceive private health insurance as a means to refund drug costs during illness and to simplify life, with many also recognizing it as a source of free major surgeries, though fewer associate it with compensation for adverse events. Additionally, over half of the respondents affirm knowledge of private health insurance, while the rest report no awareness

Low levels of enrollment in voluntary health insurance programs have been attributed, in the context of inadequate income nations, to low awareness and knowledge of the concept of health insurance. The focus of sensitization and awareness raising for health insurance activities is typically on the premium cost that prospective enrollees are anticipated to pay [14]. Health insurance has an impact on the financial cost that families must pay for care; if consumers are unaware of their insurance benefits, they may base their judgments about using medical care on erroneous estimates of the costs they would incur. Furthermore, families may get more or less insurance than is necessary if they do not comprehend their current plans (Marquis, 1983). In addition to misunderstanding the basics of health insurance, skepticism and mistrust of new health insurance systems also contribute to the decline of projects [14].

The second objective of this study is ??? to determine the private health insurance coverage among health workers. Based on findings from our study. The second question was what is the private health insurance coverage among health care providers? The results from the study shows that of the 109 participants, 96 (88.89%) have not been health insured while, 12 (11.11%) said yes they have been health insured. 105 (97.22%) said they are currently insurance whiles, 3 (2.78%) are not health insurance. 105 (97.22%) are willing to join health insurance whiles, 3 (2.78%) are not willing to join health insurance.

In many low- and middle-income nations, the private sector is the main supplier of primary healthcare for the underprivileged??. Both official, or by legally recognized regulatory authorities, and informal, or not recognized by the law, private healthcare providers are possible. For-profit and nonprofit hospitals are included in the category of formal private healthcare providers. For-profit/not-for-profit dichotomy is not so distinct in practice. Informal allopathic providers includes quacks, lay health workers, drug sellers and ordinary shop keepers. Public healthcare providers on the other hand, are health facilities built by government and the healthcare workers draw their salaries from government treasuries. This facility type is not intended for profit making [13]. Recent federal policies have prioritized raising cost-sharing as a way to transfer expenses on to consumers and deter further medical care usage [16]. Another study from

## 4. Conclusion

This study provides valuable insights into the knowledge, coverage, and determinants of private health insurance among healthcare workers in Sierra Leone.??? The findings highlight a significant level of awareness regarding private health insurance among the majority, with many recognizing its potential benefits such as cost refunds and improved quality of life,??? though gaps in understanding and skepticism persist. Coverage remains low, with most respondents lacking prior or current insurance, despite a strong willingness to participate in future schemes, suggesting a receptive attitude toward health insurance initiatives. The lack of clear evidence linking demographic and professional factors to insurance status indicates that other unexamined variables may play a critical role. These results underscore the need for targeted education campaigns to enhance comprehension and trust, alongside the development of accessible and affordable insurance models. The strong interest in joining health insurance offers a foundation for policymakers to build upon, potentially integrating private and public healthcare systems to improve coverage and access, aligning with global efforts to reduce financial barriers to healthcare. Further research is recommended to explore additional determinants and refine strategies for effective implementation.

## ABBREVIATIONS

UHC: Universal Health Coverage
WHO: World Health Organization
HCP: Healthcare Professional
CBHI: Community-based health insurance U
NIMAK: University of Makeni
ILO: International Labor Organization
SHI: Social Health Insurance
PHI: Private Health Insurance
LMIC: Low and middle-income countries
OPP: Out-of-pocket payment
LDC: Less Developed Countries
FHCI: Free Healthcare Initiative
PBF: Performance-based Financing
ACA: Affordable Care Act (ACA)

## ACKNOWLEDGEMENT

We thank the data collection team for their well-done job, the Sierra Leone Ministry of Health and Sanitation for giving access to their facility and data for this research study, and the management and administration of the University of Makeni (UNIMAK) for the provision of students who helped with the data collection process.

## DATA AVAILABILITY

All data for our research study are presented within this article and are available separately from the Lead Author or Corresponding Author with a written or electronic request from any interested individual.

## ETHICS APPROVAL and CONSENT to PARTICIPATE

Ethical approval was sought and received from the Ethical Review Board of the Sierra Leone Ethics and Scientific Review Committee. An inform consent form was issued to each respondent to seek their own knowledge, assure them that their contribution to the study will be confidential, and the data provided can only be used for the purpose of the study. The respondents were also informed that no form of any financial incentive will be given to them for being part of the study – and that their choice of not being part of the study cannot stop them from getting any benefit that may come because of the study.

## TRANSPARENCY

The authors volunteer that *chat GPT*, or any equivalent AI program, was not a source of data or information and was not used to draft or embellish our manuscript.

## AUTHORS CONTRIBUTION

**Sallu Nfagie Kamara:** Conceptualization, Investigation, Initial Data Analysis, and Formalizing Original manuscript.

**Abraham Isiaka Jimmy:** Co-Conceptualization, Data Curation, and Writing –Drafting Original Manuscript and Reviewing & Editing.

**Aiah Lebbie:** Supervision, Methodology, and Data Management.

**Lee Presley Gary Jr**.: Resources, Visualization, and Writing – Restructuring, Reviewing & Editing Final manuscript.

## BIOGRAPHIES

**Sallu Nfagie Kamara** holds a Bachelor of Pharmacy (Honors) degree from the University of Sierra Leone (Freetown), a Master Degree in Drug Discovery and Development from the University of Lagos (Nigeria), and a Master of Public Health from the University of Maken (Sierra Leone). C urrently he is a Lecturer on the Faculty of Pharmaceutical Sciences and Department of Pharmacology at the College of Medicine and Allied Health Sciences in the University of Sierra Leone, where he has been actively involved in pharmaceutical education and clinical training for o v e r six years. Mr. Kamara’s academic and research interests lie at the intersection of pharmacy, public health policy, and health systems strengthening with an extended commitment to addressing systemic gaps in healthcare financing and access for developing healthcare professionals. He is particularly focused on promoting evidence-based strategies to improve the welfare of the healthcare workforce.

**Abraham Isiaka Jimmy** is a Public Health Specialist and Researcher and University Instructor. He has more than six years of experience working in emergency management and community development contexts in Sierra Leone, while focusing on improving health program management. He teaches public health and mental health courses at the University of Makeni, located in the City of Maken, Sierra Leone. He advises on the operationalization of One Health Programs, including developing policies, strategies, and governance for mitigating public health challenges and identifying evidence-based solutions.

**Aiah Lebbie** is a pediatric surgeon with broad academic and public health backgrounds. His primary research focus is advancing and promoting pediatric surgery. Dr Lebbie is currently the Head of the Department of Surgery at the University of Sierra Leone/Teaching Hospitals Complex, located in Connaught, Freetown, Sierra Leone, and Head of the Department of Surgery at the College of Medicine and Allied Health Sciences (COMAHS) in Freetown. For over a decade, he has been actively involved in planning, designing and managing the training of medical students and specialist surgeons.

**Lee P. Gary, Jr**. is a Visiting Fulbright Research Scholar in the Department of Public Health at the University of Malta for 2025-2026. During 2024, he was a Visiting Research Scholar at the University of Makeni, located in the City of Makeni, Sierra Leone, and he is Owner/CEO of Strategic Management Services–USA, a global consultancy specializing in mitigating public health threats and hazards associated with untreated wastewater. He is a member of the Water Environment Federation and is an adjunct instructor for the National Disaster & Emergency Management University (Emmitsburg, Maryland, USA).

## RESEARCH FIELDS

**Sallu Nfagie Kamara:** Health System and Policy Research (public health financing & insurance), Ethnopharmacology (discovery of natural products), Drugs, Pharmacovigilance and rational drug use, Drug regulation, Occupational health and safety, Supply chain management

**Abraham Isiaka Jimmy:** Public Health Policy, Health System Financing, Healthcare Economics, Public Mental Health Policy, Maternal Child Health, Occupational Health Safety & Regulations, and Environmental Health Risks & Assessments.

**Aiah Lebbie:** Maternal Healthcare, Occupational Health Safety & Regulations, Environmental Health Risk & Assessments, Public Health Policy, Infectious Diseases, and Nursing Profession Education & Career Development.

**Lee Presley Gary, Jr**.: Disaster Management, Emergency Management, and Mitigation of Dirty Water and Human Waste, and Professional Development of Healthcare Workforce.

